# Colchicine use in patients with COVID-19: a systematic review and meta-analysis

**DOI:** 10.1101/2021.02.02.21250960

**Authors:** Leonard Chiu, Chun-Han Lo, Max Shen, Nicholas Chiu, Rahul Aggarwal, Jihui Lee, Young-Geun Choi, Henry Lam, Elizabeth Horn Prsic, Ronald Chow, Hyun Joon Shin

**Affiliations:** Columbia University Vagelos College of Physicians and Surgeons, Columbia University, New York City, NY, United States of America; Massachusetts General Hospital, Harvard Medical School, Harvard University, Boston, MA, United States of America; Beth Israel Deaconess Medical Center, Harvard Medical School, Harvard University, Boston, MA, United States of America; Weill Cornell Medicine, New York, NY, United States of America; Sookmyung Women’s University, Seoul, Korea; Library Services, Sunnybrook Health Sciences Centre, Toronto, ON, Canada; Yale New Haven Health, Yale School of Medicine, Yale University, New Haven, CT, United States of America; Yale School of Public Health, Yale University, New Haven, CT, United States of America; Hanyang Impact Science Research Center, Seoul, Korea; Lemuel Shattuck Hospital, Massachusetts Department of Public Health, Jamaica Plain, Boston, MA, United States of America; Brigham and Women’s Hospital, Boston, MA, United States of America

**Keywords:** colchicine, COVID-19, mortality

## Abstract

**Introduction:** Colchicine may inhibit inflammasome signaling and reduce proinflammatory cytokines, a purported mechanism of COVID-19 pneumonia. The aim of this systematic review and meta-analysis is to report on the state of the current literature on the use of colchicine in COVID-19 and to investigate the reported clinical outcomes in COVID-19 patients by colchicine usage.

**Methods:** The literature was searched from January 2019 through January 28, 2021. References were screened to identify studies that reported the effect of colchicine usage on COVID-19 outcomes including mortality, intensive care unit (ICU) admissions, or mechanical ventilation. Studies were meta-analyzed for mortality by the subgroup of trial design (RCT vs observational) and ICU status. Studies reporting an risk ratio (RR), odds ratio (OR) and hazard ratio (HR) were analyzed separately.

**Results:** Eight studies, reporting on 16,248 patients, were included in this review. The Recovery trial reported equivalent mortality between colchicine and non-colchicine users. Across the other studies, patients who received colchicine had a lower risk of mortality - HR of 0.25 (95% CI: 0.09, 0.66) and OR of 0.22 (95% CI: 0.09, 0.57). There was no statistical difference in risk of ICU admissions between patients with COVID-19 who received colchicine and those who did not – OR of 0.26 (95% CI: 0.06, 1.09).

**Conclusion:** Colchicine may reduce the risk of mortality in individuals with COVID-19. Further prospective investigation may further determine the efficacy of colchicine as treatment in COVID-19 patients in various care settings of the disease, including post-hospitalization and long-term care.

## INTRODUCTION

Coronavirus disease 2019 (COVID-19) caused by severe acute respiratory syndrome coronavirus 2 (SARS-CoV-2) is an ongoing global pandemic. Limited therapies have shown efficacy in the treatment of COVID-19. Remdesivir is recommended for use in hospitalized patients requiring supplemental oxygen, though is not routinely recommended for those who require mechanical ventilation (1–4). Dexamethasone is also recommended for these hospitalized patients, as well as those on high-flow oxygen, non-invasive ventilation, mechanical ventilation, and extracorporeal membrane oxygenation (5–7). Monoclonal antibody therapies Bamlanivimab and Casirivimab-Imdevimab, have received emergency use authorization from the United States Food and Drug Administration for non-hospitalized patients who have mild to moderate COVID-19 and certain risk factors for severe disease (8). Nevertheless, there is limited pharmacologic treatment for patients prior to hospitalization with non-severe disease.

The pathophysiology underlying severe COVID-19 pneumonia is an exaggerated inflammatory response, with an overproduction of early response proinflammatory cytokines, including tumor necrosis factor (TNF), IL-6 and IL-1β (9,10). In addition, severe acute respiratory syndrome coronavirus 1 (SARS-CoV-1), which is closely related to SARS-CoV-2, has been shown to activate the NLRP3 inflammasome (11). Thus, it has been hypothesized that therapies that present potent anti-inflammatory action and target inflammasome action may be effective therapies. Colchicine is routinely used in the treatment of inflammatory conditions such as gout, rheumatic disease, and pericarditis. Its mechanism of action is through inhibition of neutrophil chemotaxis and activity in response to vascular injury. Additionally, colchicine inhibits inflammasome signaling and reduces the production of active IL-1β (12).

Several studies have reported on the use of colchicine amongst COVID-19 patients. The COLCORONA investigators reported a randomized controlled trial of 4488 patients non-hospitalized patients with COVID-19, randomly assigning patients to colchicine or placebo for 30 days (13). In a pre-specified analysis of patients with PCR-proven COVID-19, colchicine reduced the incidence of their primary outcome—a composite of death or hospitalization— compared with placebo (OR=0.75, 0.57-0.99; p=0.04). Earlier in the pandemic when PCR reagent was lacking, clinically diagnosed COVID-19 patients were included in the analysis. In this population, the primary endpoint did not reach statistical significance (OR=0.79, 0.61-1.03; p=0.08) (13). Other randomized controlled trials (RCT) and observational studies have also reported benefits for colchicine in COVID-19 (14–18, 20–21). To our knowledge, only one prior meta-analysis has been conducted for colchicine in COVID-19 (22). This meta-analysis had several limitations, namely inclusion of smaller studies but exclusion of recent larger trials, inclusion of observational studies that reported unadjusted estimates of benefit, and lack of stratified analyses by study design.

The aim of this systematic review and meta-analysis is to report on the state of the current literature on the use of colchicine for COVID-19, and to investigate the reported clinical outcomes in COVID-19 patients by colchicine usage.

## METHODS

### Search Strategy

The databases of Ovid MEDLINE, Embase, the Cochrane Central Register of Controlled Trials medRxiv, and researchsquare.com were searched from January 2019 through March 24, 2021. The search strategy is presented in Appendix 1.

### Study Screening

Two reviewers (LC, C-HL) independently assessed articles through level 1 title and abstract screening, and level 2 full-text screening. In the case of discrepancies, a discussion occurred between the two reviewers and consensus was achieved on whether to include or exclude a particular study. If consensus could not be achieved, a third reviewer (RC) was consulted.

Articles were eligible for inclusion after level 1 screening if the title and/or abstract mentioned COVID-19 and colchicine. These articles then underwent level 2 screening, at which point the full text was reviewed to identify primary research articles that reported COVID-19 outcomes from a clinical or claims database.

### Quantitative Synthesis

Articles were included for quantitative synthesis if they (1) were of observational study design and reported a relative risk measure adjusted for possible confounders, or (2) were of RCT and reported either a relative risk measure or event data, and the article compared colchicine recipients to non-colchicine recipients with respect to the outcomes of mortality, mechanical ventilation, ICU admission, or hospital length of study. Study demographics (i.e. central measure of tendency for age, percentage male) were also noted for each included study.

As with study screening, this process was done by two reviewers (LC, C-HL), and a third reviewer (RC) was consulted if consensus could not be achieved.

### Risk of Bias Assessment

Risk of bias was assessed using tools developed by the Cochrane Bias Methods Group, with the Risk Of Bias In Non-randomized Studies – of Interventions (ROBINS-I) tool used for the observational studies and a revised Cochrane risk of bias tool for randomized trials (RoB 2 tool) used for randomized controlled trials (23, 24).

### Statistical Analyses

Studies were meta-analyzed for each outcome and by the subgroups of colchicine use relative to hospitalization (non-hospitalized patients, hospitalized non-ICU patients, and hospitalized ICU patients), study design (RCT and observational study), and ICU status (ICU patients and non-ICU patients). Studies were meta-analyzed together according to their relative risk ratio; studies reporting on odds ratio (OR) were meta-analyzed to produce a summary OR, and studies reporting a hazard ratio (HR) were separately meta-analyzed to produce a summary HR. For RCTs which reported exclusively event data, ORs were calculated using the Haldane-Anscombe correction. In the event that the meta-analysis’s heterogeneity of I^2^ was equal to or greater than 50%, a random-effects DerSimonian-Laird analysis model was applied; otherwise (i.e. when there was heterogeneity of I^2^ less than 50%), a fixed-effects inverse-variance analysis model was used. A *p*-value of less than 0.05 was considered statistically significant in the test for overall effect.

Publication bias was assessed visually, using a funnel plot, and statistically, using Egger’s test. A *p*-value of > 0.05 indicated low concern for publication bias. All analyses were conducted using Stata 17.

### Patient and Public Involvement

Patients or the public were not involved in the design, or conduct, or reporting, or dissemination plans of our research. This systematic review and meta-analysis did not require an IRB submission as data is all publicly available online.

## RESULTS

Of 540 records that underwent level 1 screening, 537 were identified through database search and 3 identified through backward reference searching. After removing duplicates, 417 records underwent level 1 screening. Subsequently, 129 articles underwent level 2 screening. Ten studies were assessed for quantitative synthesis, of which 8 reported adjusted relative risk ratios and were included in this systematic review and meta-analysis (13, 15–21) (Appendix 2).

Study demographics are presented in Table 1. Five studies were RCTs and three studies were observational retrospective cohort studies. While Rodriguez-Nava *et al* reported the effects of colchicine on ICU patients, the other seven studies reported on non-ICU patients. Tardif *et al*, defined a colchicine user as one who used colchicine after COVID-19 diagnosis but before a COVID-19 hospitalization, while the other seven studies defined colchicine users as ones who used colchicine after a COVID-19 hospitalization.

**Table 1.**
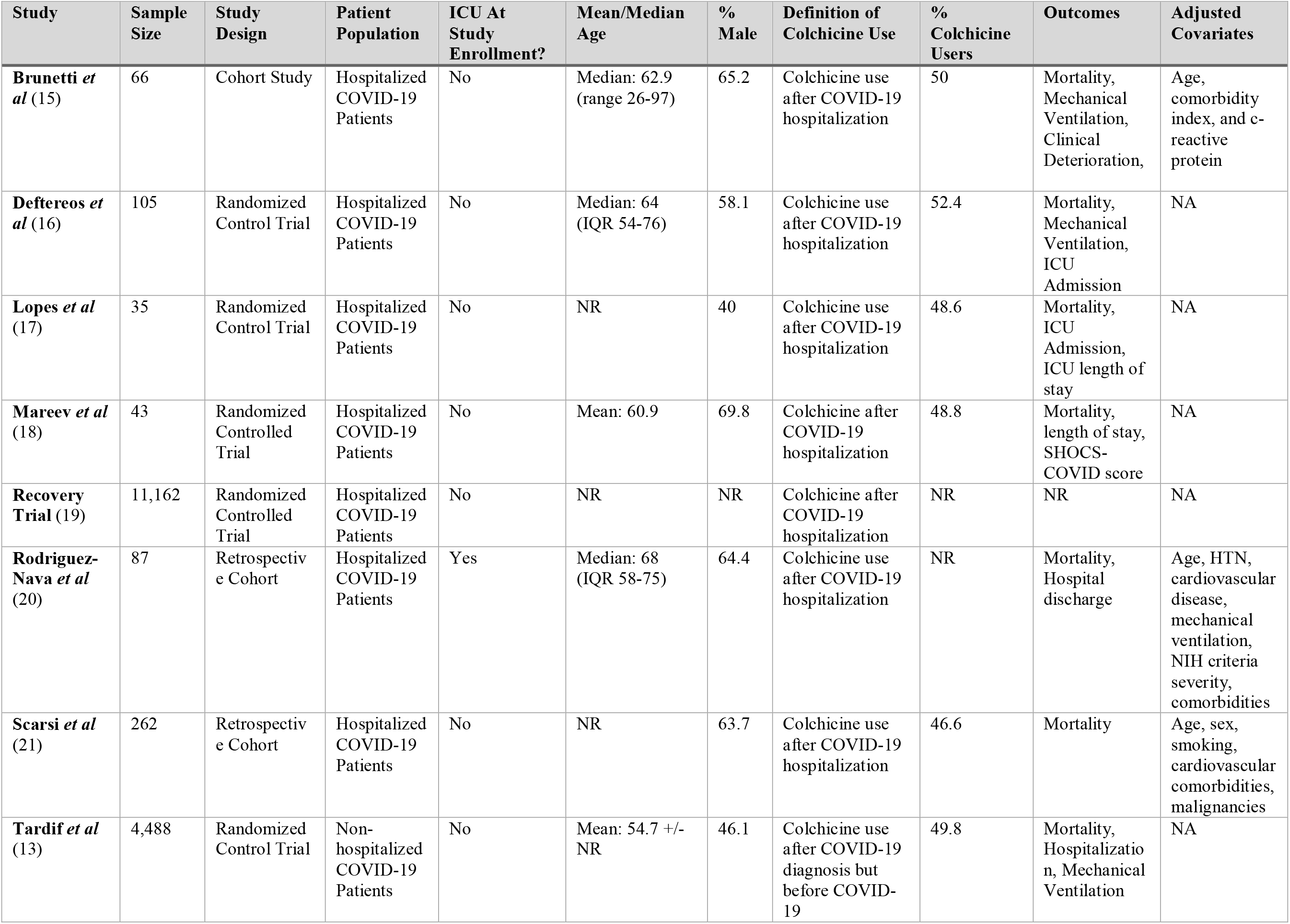
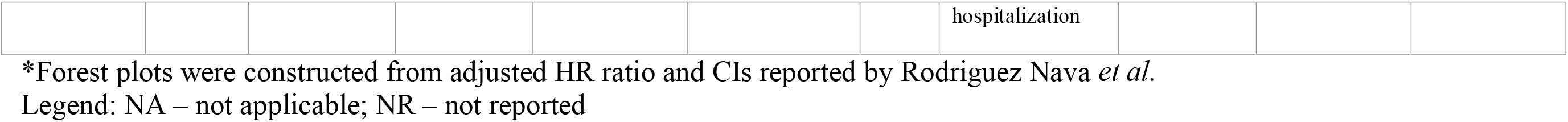
Study Demographics

Risk of bias assessment is presented in Appendix 3. The risk of bias of the observational studies is presented in Appendix 3a and the risk of bias of the RCTs is presented in Appendix 3b. Visual inspection of the funnel plot and the p-value of p=0.03 Egger’s test indicated some concern for publication bias (Appendix 4).

### Mortality

Eight studies, reporting on 16,248 patients, compared mortality between colchicine users and non-users. According to Tardif *et al*, there is no difference in mortality risk between colchicine users and non-users prior to hospitalization for COVID-19—OR of 0.56 (95% CI: 0.19, 1.66). When colchicine is used after hospitalization and excluding the Recovery trial, patients who received colchicine had a lower risk of mortality - HR of 0.25 (95% CI: 0.09, 0.66) and OR of 0.22 (95% CI: 0.09, 0.57). The Recovery trial reports no difference in mortality—OR of 1.02 (95% CI: 0.94, 1.11) (Figure 1a).

**Figure 1.**
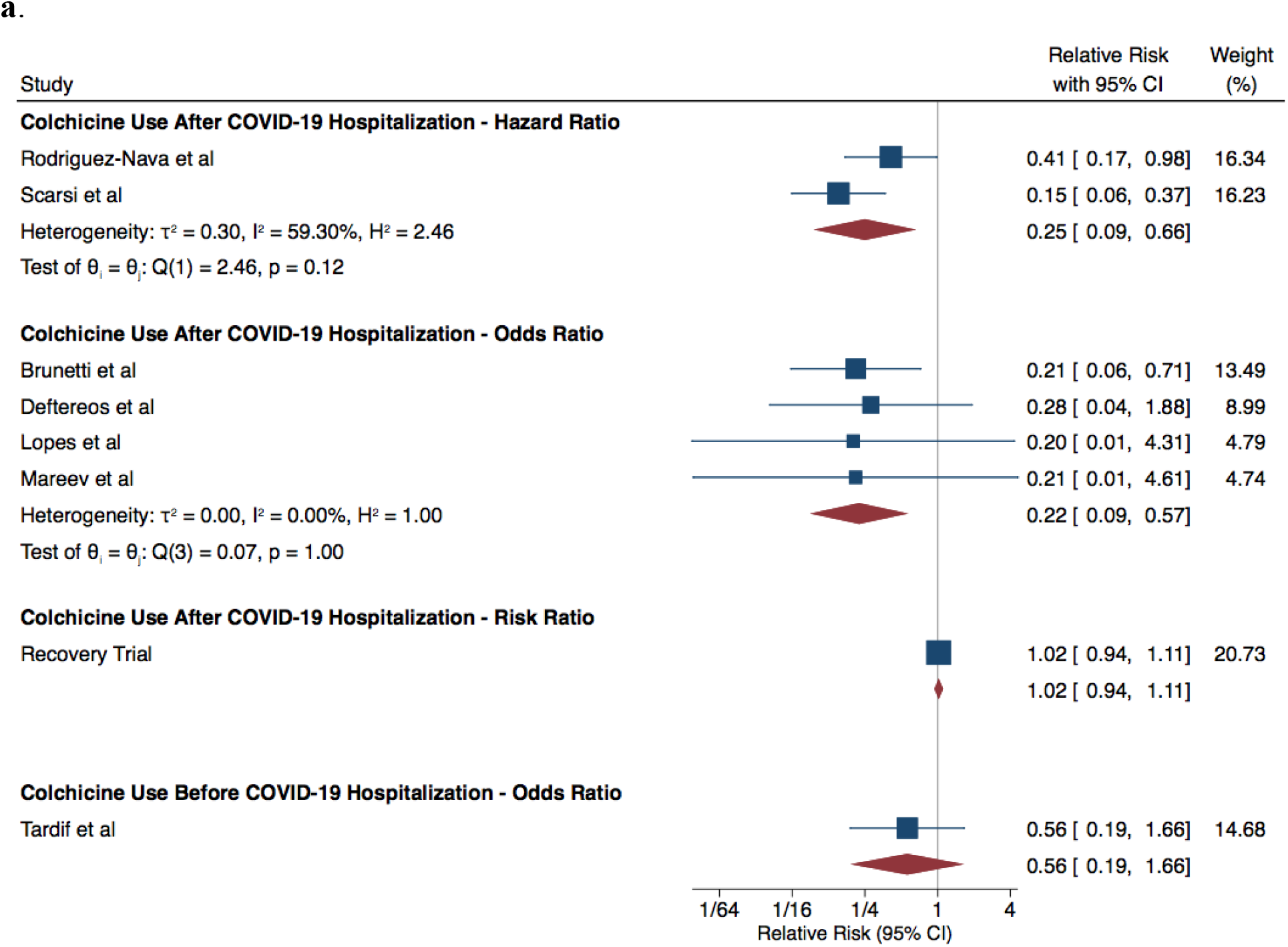
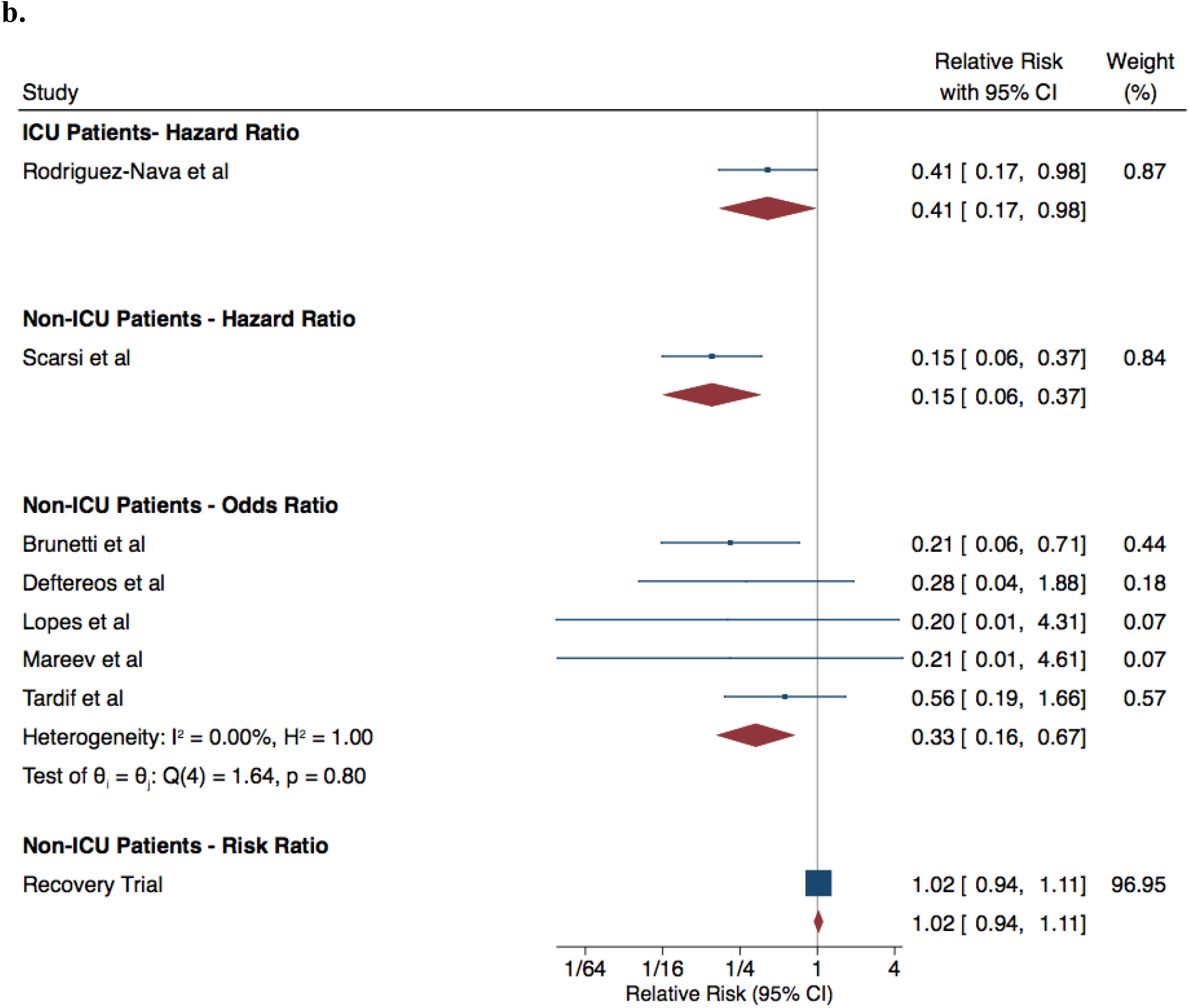
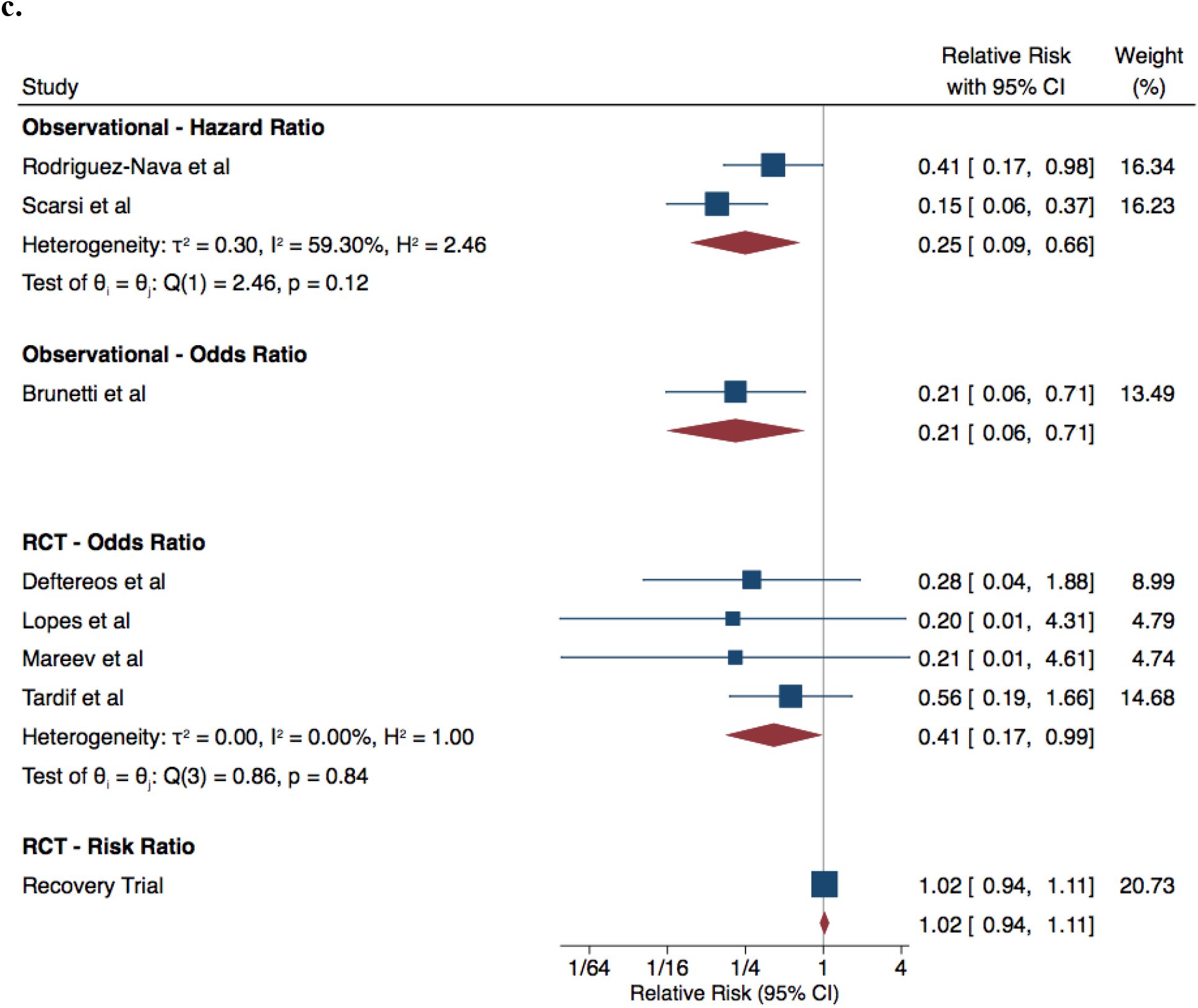
COVID-19 Patients Who Used Colchicine vs Did Not Use Colchicine – Mortality Analysis **1a** By Colchicine Use **1b** By Timing of Colchicine Use Relative to Hospitalization **1c** By Study Design

Subgroup analyses by ICU status and study design are presented in Figures 1b and 1c. Excluding the Recovery trial, colchicine seems to be associated with a lower risk of mortality among ICU patients, non-ICU patients, in observational studies and in randomized controlled trials.

### ICU Admissions

Two studies reported the risk of ICU admissions among patients with COVID-19 and colchicine use. There was no statistical difference in risk of ICU admissions between patients with COVID-19 who received colchicine and those who did not – OR of 0.26 (95% CI: 0.06, 1.09) (Figure 2).

**Figure 2.**
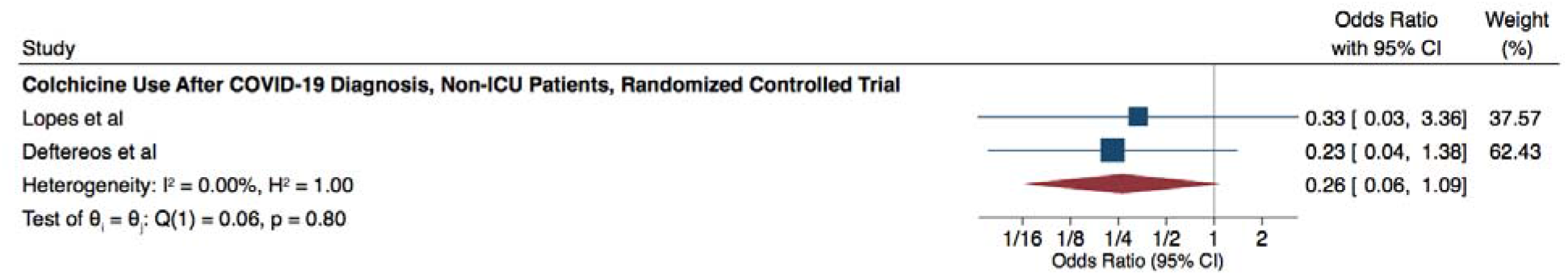
COVID-19 Patients Who Used Colchicine vs Did Not Use Colchicine – ICU Admission Analysis

### Mechanical Ventilation

Two studies reported on the risk of mechanical ventilation among patients with COVID-19 and colchicine use. Tardif *et al* reported that among patients diagnosed with COVID-19 who used colchicine before hospitalization, there was no significant difference in risk of mechanical ventilation between patients who received colchicine and those who did not – OR: 0.53 (95% CI: 0.25, 1.09). Deftereos *et al* reported that among patients diagnosed with COVID-19 who used colchicine after hospitalization, there was no significant difference in risk of mechanical ventilation between patients who used colchicine and those who did not – OR: 0.90 (95% CI: 0.49, 1.67).

## DISCUSSION

We present the first comprehensive systematic review and meta-analysis investigating the effects of colchicine use on COVID-19 patients, meeting the methodological guidance for a high-quality systematic review and meta-analysis as recommended by the MOOSE group (23). Our findings, including eight studies, suggest that among existing studies except for the Recovery trial, patients taking colchicine after COVID-19 diagnosis had a lower risk of mortality in the pooled estimate. With eight studies reporting on 16,248 patients in total, this meta-analysis has greater statistical power than the meta-analysis of Vrachatis *et al*, which reported on six studies and 881 patients (20).

More specifically, this systematic review and meta-analysis differs from the meta-analysis by Vrachatis *et al* (20) in three ways. First, our review included results of several recent trials, including the recent COLCORONA trial with 4,488 patients, results of another recent observational study of 87 patients (17) and the Recovery trial with 11,162 patients. Second, our review included only the results of RCTs and observational studies with adjusted relative risk ratios. In particular, Vrachatis *et al* included two observational studies that did not report adjusted OR which were excluded in our analysis (24, 25). Given that covariate distributions of colchicine user and non-user groups cannot be assumed to be similar as in RCTs, excluding unadjusted relative risk ratio results in observational data is necessary to reduce confounding bias. Third, our review separated the overall mortality analysis by study design, colchicine use before and after hospitalization, and ICU status.

In the trial by Tardif *et al*, in a pre-specified analysis of patients with PCR-proven COVID-19, the colchicine group showed a reduction in the primary outcome which was a composite rate of death or hospitalization compared with placebo—OR=0.75, 95% CI: 0.57-0.99; p=0.04 (13). The pre-specified analysis of patients with PCR-proven COVID-19 excluded clinically diagnosed COVID-19 patients—a diagnostic criteria that was allowed in the initial stages of the pandemic given PCR reagent shortages. The reduction in the trial’s composite primary outcome (a composite of death and hospitalizations) was driven in large part by reduction in hospitalizations of patients with COVID-19. With a low event rate in either arm (5/2075 in colchicine arm vs. 9/2084 in the non-colchicine arm), the trial may have been underpowered to detect a difference in the outcome of death in the non-hospitalized patients with COVID-19.

Given that some studies in this analysis studied hospitalized patients and others studied non-hospitalized patients, it is informative to identify the timing of the exposure of colchicine to patients in relation to the timing of their hospitalization if possible (Figure 3). All studies in this meta-analysis were exposed to colchicine after COVID-19 diagnosis and seven of the eight studies included patients exposed to colchicine after hospitalization. The trial by Tardif *et al* found that colchicine is beneficial in non-hospitalized patient in terms of preventing hospitalization or death. Without the Recovery trial, our results suggested that among non-ICU hospitalized patients, colchicine users were at a lower risk of mortality relative to non-colchicine users. Rodriguez-Nava *et al*, reported that ICU patients had a lower risk of mortality when using colchicine.

**Figure 3.**
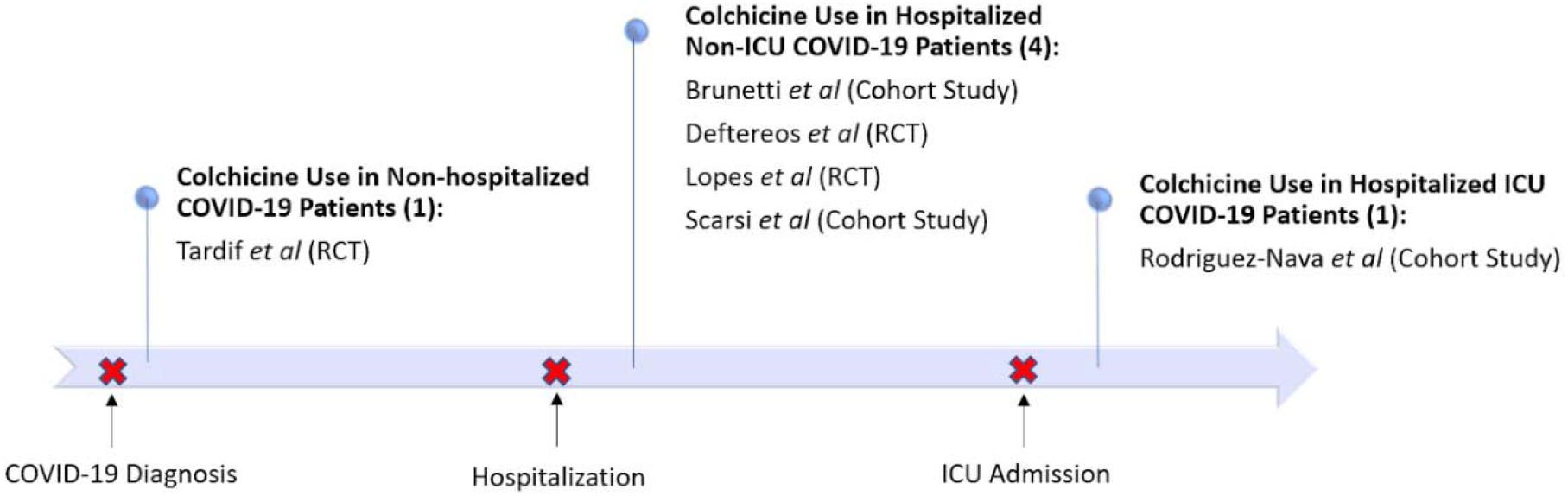
Studies Examining Colchicine at Various Care Settings

However, the Recovery trial (19) reports a different result – colchicine and non-colchicine users have equivalent, not reduced, risk for mortality risk. This trial has been stopped early by their Data Monitoring Committee, and limited data is currently available regarding the efficacy of colchicine among COVID-19 patients. We await the publication of the complete results of the Recovery trial. It is important to note that trials that undergo early termination are typically one of two types – reporting significant benefit or significant harm of the intervention, relative to its comparator; studies that undergo early termination therefore are more likely to suffer from biases away from the null and true effect estimate. Nevertheless, the Recovery trial should serve as a cautionary tale for further research in colchicine, despite the reported superiority of colchicine across all other published data.

If there is any solace in these results, it is that colchicine may be beneficial or have a null effect in patients with COVID-19. Given that colchicine is generally a well-tolerated drug, is a relatively inexpensive drug, and, despite known diarrhea rates of 23% in colchicine users (26), has a good safety profile as shown in a meta-analysis on the adverse effects of colchicine (27) and in Tardif *et al*’s study (13), there is potential for further investigation in the use of colchicine for COVID-19 patients. Further large, well-powered randomized studies could be of value to ascertain the effect colchicine would have on both mortality and other clinically meaningful endpoints such as hospitalization, ICU admission, and need for mechanical ventilation in the three different categories of illness: 1) non-hospitalized patients, 2) hospitalized patients, and 3) patients in ICUs (Figure 3).

This study is not without limitations. Intrinsic to meta-analysis study designs, the strength of meta-analysis conclusions is limited to the strength of the input studies and underlying data. This meta-analysis contains a mix of observational and RCT data. To reduce bias in grouping observational and RCT, studies were grouped by trial design when comparing mortality in colchicine and non-colchicine users and only studies reporting adjusted risk estimate were used in observational studies. Additionally, all included studies had some concern for risk of bias. Moreover, this review only contains eight studies, with only two studies examining the risk of ICU admission and two evaluating the risk of mechanical ventilation. Furthermore, the long-term effects of COVID-19 are a significant clinical concern and a major source of disability and health care utilization. Further studies in post-hospitalization and long-term care in patients with COVID-19 are also warranted.

In conclusion, our meta-analysis suggests that colchicine may reduce or lead to equivalent risk of mortality in individuals with COVID-19, based on the existing observational studies and trials. Further prospective investigation may further determine the efficacy of colchicine as treatment in COVID-19 patients in various care settings of the disease, including post-hospitalization and long-term care.

## Data Availability

N/A

## Acknowledgements

Authors have no funding to disclose.

## Appendix 1. Search Strategies

Database: Ovid MEDLINE(R) and Epub Ahead of Print, In-Process & Other Non-Indexed Citations, Daily and Versions(R) <1946 to March 24, 2021>

Search Strategy:

--------------------------------------------------------------------------------

1. (covid 19 or covid-19).mp. (95589)
2. “coronavirus disease 2019”.mp. (18596) 3 SARS-CoV-2.mp. (37725)
3. or/1-3 (99157)
4. exp Colchicine/ (15163)
5. colchicine.mp. (20749)
6. colcrys.mp. (8)
7. mitigare.mp. (4)
8. gloperba.mp. (0)
9. or/5-9 (21710)
10. 4 and 10 (90)
11. 12 limit 11 to english language (88)

***************************

Database: Embase Classic+Embase <1947 to 2021 March 24>

Search Strategy:

--------------------------------------------------------------------------------

1. (covid 19 or covid-19).mp. (80912)
2. “coronavirus disease 2019”.mp. (81124) 3 SARS-CoV-2.mp. (28874)
3. or/1-3 (93686)
4. exp Colchicine/ (35809)
5. colchicine.mp. (40067)
6. colcrys.mp. (73)
7. mitigare.mp. (2)
8. gloperba.mp. (0)
9. or/5-9 (40067)
10. 4 and 10 (207)
11. 12 limit 11 to english language (205)

***************************

Database: EBM Reviews - Cochrane Central Register of Controlled Trials <February 2021>

Search Strategy:

--------------------------------------------------------------------------------

1. (covid 19 or covid-19).mp. (3627)
2. “coronavirus disease 2019”.mp. (707)
3. SARS-CoV-2.mp. (1379)
4. or/1-3 (3725)
5. exp Colchicine/ (337)
6. colchicine.mp. (930)
7. colcrys.mp. (2)
8. mitigare.mp. (0)
9. gloperba.mp. (0)
10. or/5-9 (931)
11. 4 and 10 (40)
12. limit 11 to english language (7)

***************************

Database: medRxiv – The Preprint Server for Health Sciences <January 28, 2021>

--------------------------------------------------------------------------------

1. covid 19 AND (colchicine OR colcrys OR mitigare OR gloperba) (52)

***************************

Database: Research Square Preprint Platform <January 28, 2021>

--------------------------------------------------------------------------------

1. covid 19 AND (colchicine OR colcrys OR mitigare OR gloperba) (2)

## Appendix 2. PRISMA Flow Diagram

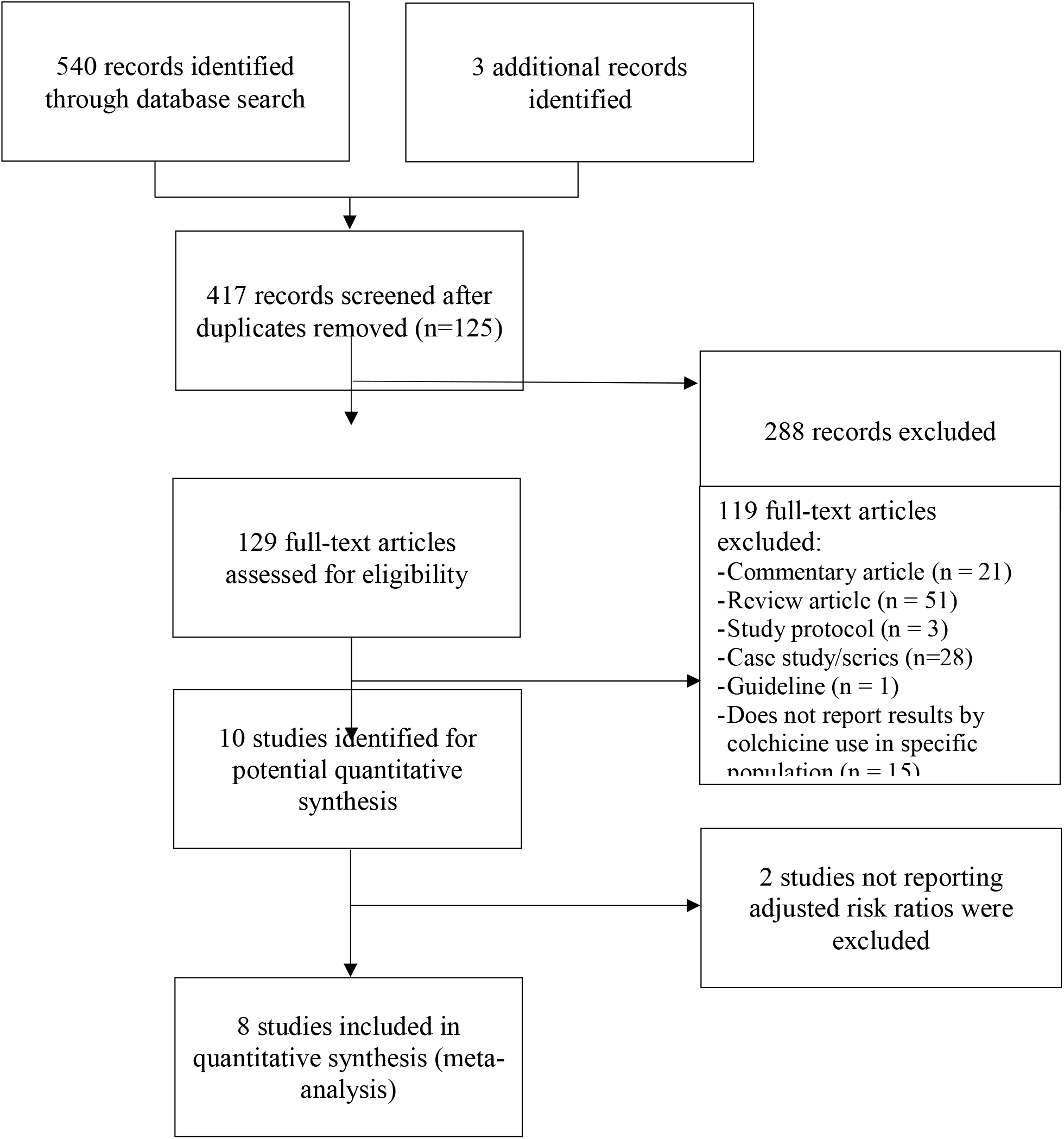

## Appendix 3. Risk of Bias Assessment 3a Observational Studies 3b Randomized Controlled Trials

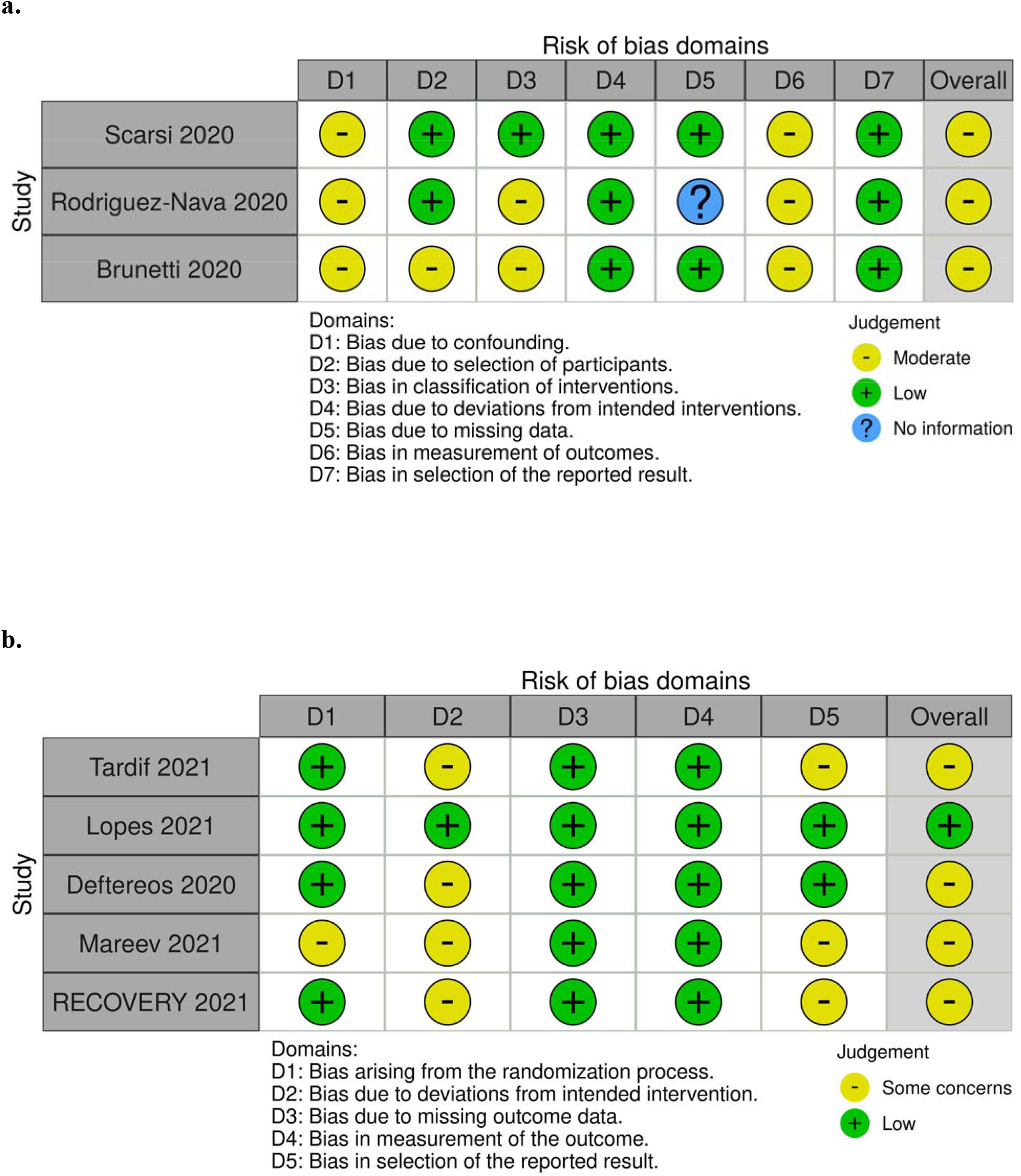

## Appendix 4. Assessment for Publication Bias – Funnel Plot

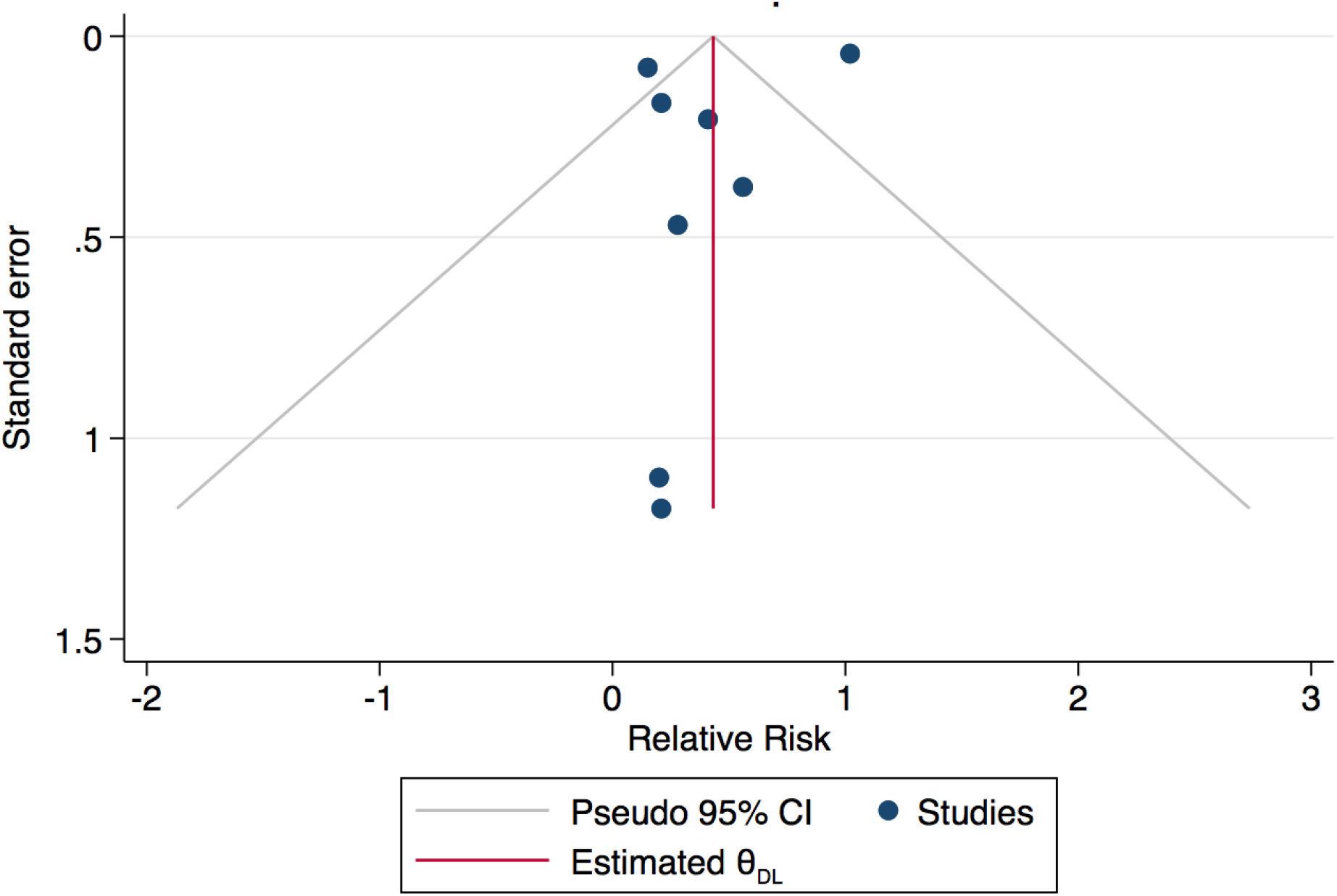

